# LWU-Net approach for Efficient Gastro-Intestinal Tract Image Segmentation in Resource-Constrained Environments

**DOI:** 10.1101/2023.12.05.23299425

**Authors:** Marreddi Jayanth Sai, Narinder Singh Punn

## Abstract

This paper introduces a Lightweight U-Net (LWU-Net) method for efficient gastro-intestinal tract segmentation in resource-constrained environments. The proposed model seeks to strike a balance between computational efficiency, memory efficiency, and segmentation accuracy. The model achieves competitive performance while reducing computational power needed with improvements including depth-wise separable convolutions and optimised network depth. The evaluation is conducted using data from a Kaggle competition-UW Madison gastrointestinal tract image segmentation, demonstrating the model’s effectiveness and generalizability. The findings demonstrate that the LWU-Net model has encouraging promise for precise medical diagnoses in resource-constrained settings, enabling effective image segmentation with slightly less than a fifth of as many trainable parameters as the U-Net model.

## 1. Introduction

In 2019, approximately 5 million individuals were diagnosed with gastro-intestinal (GI) tract malignancies, thereby exacerbating the global epidemiological load of this pathology. Of this cohort, nearly 50% were identified as viable candidates for radiotherapy, a pivotal modality in oncological management. Nonetheless, radiation oncologists encounter a formidable challenge in administering precise radiation dosimetry while concurrently safeguarding adjacent critical structures, such as the gastric and intestinal regions. Utilizing MR-Linacs, which amalgamate magnetic resonance imaging (MRI) with linear accelerator modalities, has indeed heralded a paradigm shift in radiation therapy. However, the manual demarcation of organ contours persists as a laborious and time-intensive procedure. This not only exacerbates treatment durations but also intensifies the physical and psychological effects experienced by patients already contending with the ramifications of their pathology.

Medical image segmentation is a critical step in many diagnostic and therapeutic procedures. Over the last decade, the evolution of deep learning techniques has greatly advanced this field, resulting in improved accuracy and efficiency in image analysis. The segmentation of GI tract is vital for numerous clinical applications, including the diagnosis of cancers, inflammation, and other pathologies, as well as for the planning and monitoring of therapeutic interventions(Gilja et al., 2007). Precise segmentation of the GI tract is therefore not just a computational challenge but a necessity for improving patient care. In the segmentation of various medical images for different applications, the U-Net architecture has exhibited a notably high success rate. Given this success, numerous U-Net variants have emerged to address specific challenges inherent to biomedical image segmentation (Punn and Agarwal, 2022c).

While the conventional U-Net and its numerous derivatives have been successful in many medical imaging contexts, their utility in real-time applications or environments with computational constraints remains an issue. The traditional U-Net, despite its strengths, relies on a deep architecture that involves a substantial number of trainable parameters (Falk et al., 2019). Although this depth and complexity have contributed to its success in diverse medical imaging tasks, they also add to its computational demands. Various adaptations of the U-Net have been proposed to improve its performance or extend its applicability. Still, the challenge of efficiency, especially in resource-constrained contexts, remains relatively unaddressed (Litjens et al., 2017). The computational intensity of these architectures, coupled with the growing need to process vast quantities of medical imaging data promptly, has necessitated the development of more efficient models.

In light of the previously mentioned challenges, the Lightweight U-Net (LWU-Net) model has been introduced. LWU-Net offers a computer-friendly option that aims to balance computing power with accurate image segmentation. Although primarily designed for gastro-intestinal tract images, the concepts behind LWU-Net might be applied to other types of medical imaging as well. The present research aims to analyse the effectiveness of the proposed LWU-Net model for GI tract image segmentation and evaluate its performance against the various U-Net models. The key focus of this paper is as follows:

- The proposed model offers a captivating balance between computational efficiency and segmentation accuracy for GI tract Image Segmentation.
- Leveraging the potential of depthwise separable convolutions, preserving the capability to delineate the boundaries accurately and reducing the computational power needed.
- Conducting an evaluation and comparison of the proposed LWU-Net architecture against the U-Net based models.

The rest of the paper structure is organized as follows: Section 2 reviews the existing literature on GI tract image segmentation and LWU-Net models. Section 3 details the methodology, including the dataset used, preprocessing techniques applied, and the specific architecture and training process of the LWU-Net model. Later, in Section 4 the results and experiments performed are demonstrated, Section 5 discusses the strengths and weaknesses of the model and future research directions. Finally, the concluding remarks are presented in Section 6.

## 2. Related work

The demand for efficient and lightweight models has grown with the increasing need for real-time applications and resource-constrained environments. Several studies have focused on developing lightweight architectures to reduce computational complexity without compromising performance. MobileNet Howard et al. (2017) and ShuffleNet Zhang et al. (2018) introduced depth-wise separable convolutions and channel shuffling techniques to reduce parameters and computational costs significantly. Agarap (2018) introduces rectified linear units as classification function. Tang et al. (2019) uses separable-UNet for efficient skin lesion segmentation and Chen et al. (2018) encorporates encoder-decoder with atrous separable convolution for semantic image segmentation.

In medical image analysis, segmentation is crucial in diagnosis, treatment planning and disease monitoring. U-Net Ronneberger et al. (2015), the foundation of our proposed architecture, has been widely adopted in medical imaging tasks due to its ability to capture detailed anatomical structures. Many studies have explored different variants of U-Net for specific medical segmentation tasks Punn and Agarwal (2022c), such as brain tumor segmentation Punn and Agarwal (2021), retinal vessel segmentation Yue et al. (2019), anatomical brain segmentation using DenseNetGottapu and Dagli (2018), Tran et al. (2021) employed a Triple-unet with multi-scale input features and dense skip connection and Huang et al. (2020) proposed a UNet3+ a full scale connected U-Net, Xiao et al. (2018) proposed a weighted ResUNet for high quality retina vessel segmentation and Punn and Agarwal (2022a) proposed a self-supervised Unet framework for biomedical image segmentation applications, Jafari et al. (2020) employed an efficient deep convolutional neural network, while Punn and Agarwal (2022b) proposed attention based U-Net and Li et al. (2020) employed a nested attention aware U-Net for liver CT image segmentation Accurate segmentation of the GI tract from medical images is essential for various applications, including polyp detection, pathology assessment, and surgical planning. Several studies have focused on GI tract segmentation, with approaches ranging from traditional techniques to deep learning-based methods.

For instance, Wang et al. (2020) employed a multi scale context guided deep network, Li and Liu (2022) employed a multi-view U-Net for GI tract segmentation, Guggari et al. (2022) proposed RU-Net for GI tract image segmentation,Zhang et al. (2017) proposed the Gastric precancerous diseases classification using CNN, Asperti and Mastronardo (2017) shows the effectiveness of data augmentation for detection of gastrointestinal diseases, Ali et al. (2021) shows detection and segmentation of artefact and disease instances in gastrointestinal endoscopy, Nemani and Vollala (2022) employed LeViT U-Net++ for GI tract and Jha et al. (2019) showed the usage of ResUnet++ architecture for polyp detection and segmentation.

Previous research have explored the application of U-Net and various deep learning architectures in segmenting gastro-intestinal (GI) tract images. However, these methodologies frequently necessitate significant computational resources, especially in constrained environments. Consequently, there emerges a compelling imperative to assess the viability of employing the Lightweight U-Net model explicitly for GI tract image segmentation. Thus, this investigation endeavors to rigorously ascertain the aptness of the lightweight U-Net model for such segmentation tasks and juxtapose its efficacy against conventional U-Net architectures.

## 3. Methodology

### 3.1. Dataset

The dataset utilized in this investigation is sourced from the UWMadison GI tract image segmentation challenge presented on Kaggle (Happyharrycn, 2022). Comprised of de-identified patient scans procured from the UWMadison Carbone Cancer Center, the dataset is intended for the semantic segmentation of GI tract imagery. Scans within this compilation span a duration of 1-5 days and are manifested as 16-bit image slices. A visual representation of this data, including two exemplar images accompanied by their respective ground truth masks, can be observed in Fig. 1.

**Figure 1:**
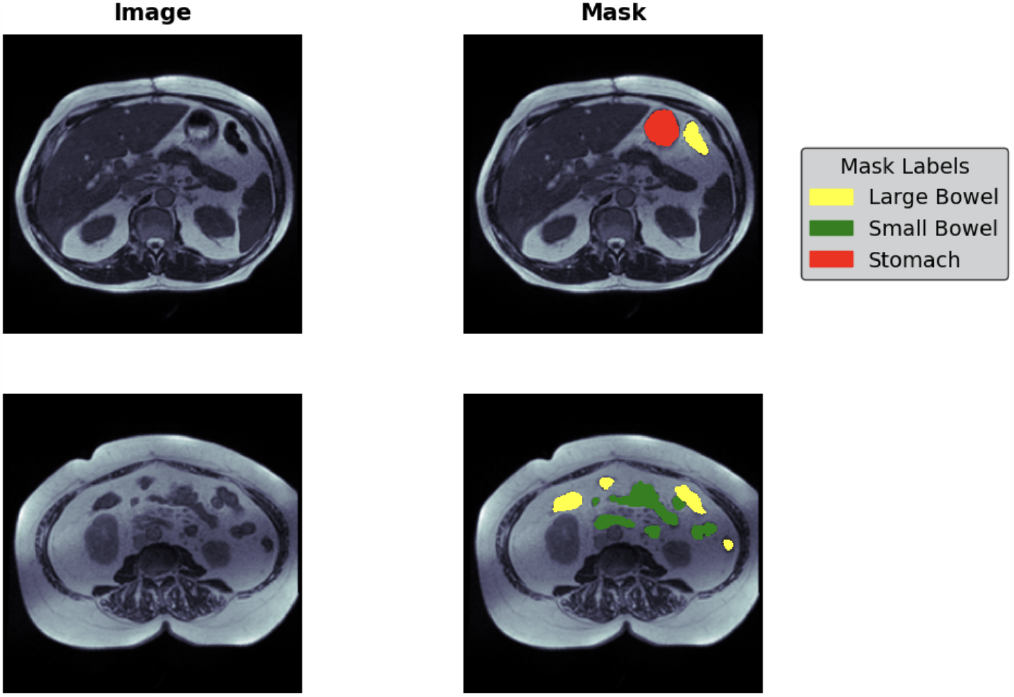
Dataset (scans with masks of organs overlapped onto the scan)

### 3.2. Preprocessing

Run-length encoding (RLE) is employed in training the provided masks, RLE is used to reduce the size of data by encoding consecutive repeated elements as a single value followed by a count of how many times that value is repeated, hence using RLE would optimise memory and bandwidth usage. One issue with these scans is the inconsistency in their widths and heights, with some images being square while others are rectangular.

To address this issue, a resizing operation is performed to standardize the dimensions of the images. Specifically, all images are resized to a fixed size of 256×256 pixels. Regarding the choice of resizing versus padding, it is noteworthy that the image size in the case of padding would be 288×288 pixels. However, it is important to consider that the majority of the pixels in these images are already black (non-informative regions), and a significant proportion of the images have empty segmentation masks. Therefore, the inclusion of padding to accommodate the maximum size is deemed unnecessary as it would introduce additional non-informative regions. Consequently, resizing the images to 256×256 pixels is deemed appropriate for this research. The images are read using OpenCV, to enhance the consistency of pixel intensity values across different images, a normalization step is applied before resizing. The resulting image is an 8-bit unsigned integer.

The dataset is then grouped based on a case ID for the sake of splitting the dataset into training and validation sets. This ensures that samples from the same case are kept together during the cross-validation process. The split is done in such a way that the distribution of images with empty segmentation is balanced across the folds. This approach helps mitigate any potential bias that may arise due to imbalanced representation of images without masks across different folds, resulting in more reliable and accurate model evaluation.

While going through the dataset, the masks for case7 day0 and case81 day30, visualised in Fig. 2, were found to be incorrect and were consequently removed from the dataset before training. These masks did not align properly with the anatomy of the GI tract, and their inclusion could have adversely affected the training process. Removing them ensured that the training dataset only contained accurate and reliable annotations, improving the quality and reliability of the model’s training process.

**Figure 2:**
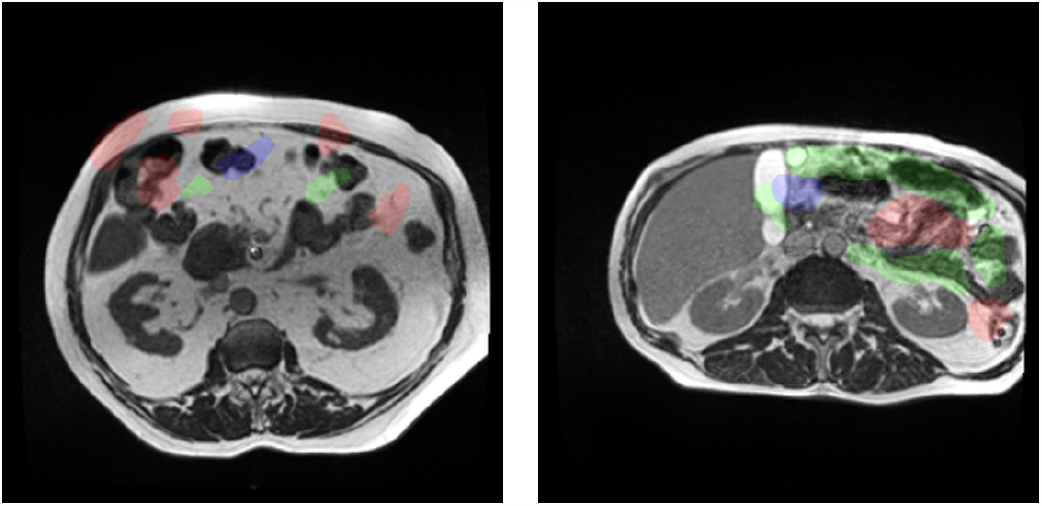
case7-day0-slice-0096 (left) and case81-day30-slice-0096 (right), shows incorrect segmentation

#### 3.3. Architecture

The LWU-Net architecture aims to decrease the computational demands of the U-Net architecture while maintaining effective segmentation capabilities. It retains the core structure of the original U-Net but replaces standard convolutions with separable convolutions. As shown in the Fig.3, the architecture consists of an encoder decoder framework with skip connections for multi scale feature fusion. The encoder part of Lightweight UNet contains a downsampling path, similar to the original U-Net. Each downsample block in the encoder includes a separable convolution layer, followed by a batch normalization layer and a ReLU activation function.

**Figure 3:**
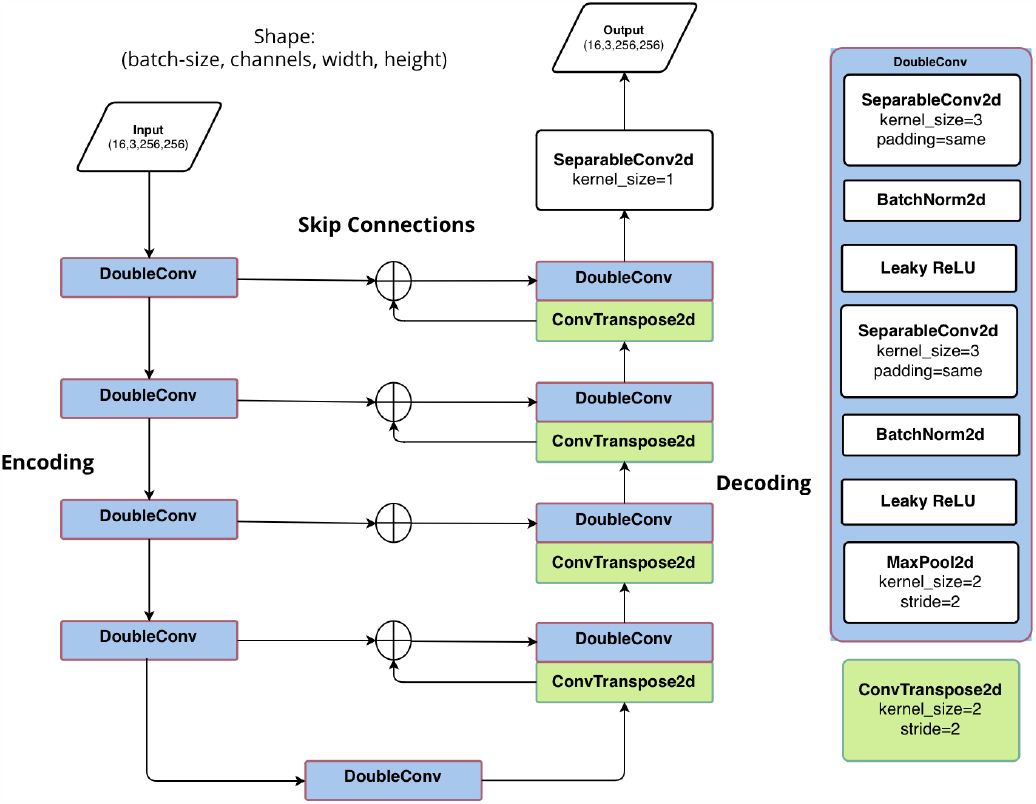
Visual Representation of LWU-Net Architecture

The downsampling utilises the separable convolution in place of the traditional convolutions, which reduces spatial dimensions while preserving crucial feature information as a separable convolution first employes the depth-wise convolution and there after applies point-wise convolution. This downsampling process gradually extracts high level features while maintaining a smaller feature map size because these are convolutions that can be decomposed into smaller convolutions along their spatial dimension. This means that a single, large convolution operation, such as the original convolution layer, can be broken down into a series of smaller convolutions that, when applied sequentially, yield the same outcome. As a result, this decomposition reduces the number of required multiplicative operations(Eq. 3) while still achieving the identical result.

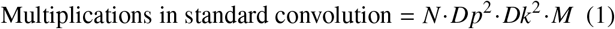

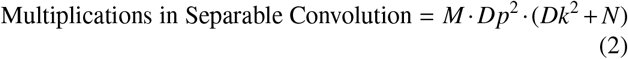

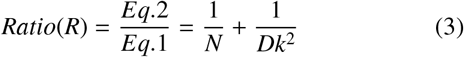

Where:

- *N* : Number of filters
- *M* : Number of channels
- *D f* : Input size
- *Dk* : Kernel size
- *Dp* : Output size

The decoder part of LWU-Net follows the standard U-Net architecture. Every upsampling block comprises a transpose layer to increase spatial dimensions. Followed by concatenation with the corresponding encoded feature maps. This skip connection allows for the fusion of low level and high level features. Enabling accurate segmentation. After concatenation, a separable convolution layer is applied to capture spatial dependencies and generate refined feature representations. To normalize the features a batch normalization layer is then applied, followed by a ReLU activation function to introduce non linearity. The final output of the decoder is a segmentation map that matches the exact spatial dimensions as the input image.

Overall, the LWU-Net architecture substitutes the traditional convolutions in the original UNet with separable convolutions. This modification produces a more lightweight model that has reduced computational complexity. By harnessing the advantages of separable convolutions, there is a notable reduction in parameters and computational operations without compromising the competitive segmentation performance as compared to other U-Net variants.

## 4. Experiments and results

The performance metrics used are Intersection over Union (Jaccard coefficient) and dice coefficient, and the loss functions are derived from the combinations of Binary Cross Entropy (BCE) Loss, Dice Loss, Focal Loss.

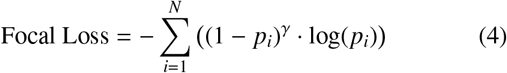

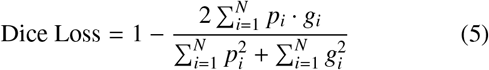

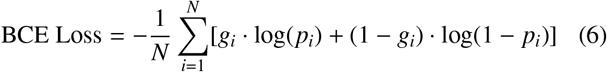

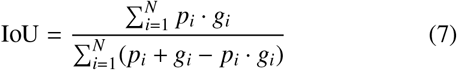

where:

- *N* is the number of pixels/elements in the segmentation mask
- *p*_*i*_ is the predicted probability for pixel/element *i*
- *g*_*i*_ is the ground truth value for pixel/element *i*
- γ is the focusing parameter that controls the degree of emphasis on hard examples.

The intial experiment with the loss function as BCE loss (Eq.6) + dice loss (Eq. 5 resulted in inferior scores when compared to the focal loss (Eq. 4) + dice loss (Eq. 5), weighted loss options like 0.9*DiceLoss + 0.1*BCE Loss were also tried but as there are significantly large amount of black pixels in the image and significant amount of images with empty segmentation masks, the bce loss or the focal loss tends to stagnate at the end of the training. These metrics were utilized to assess the performance of our model before running the predictions on Kaggle’s hidden test set. The comparitive plot of their validation dice coefficients is shown in the Fig. 4.

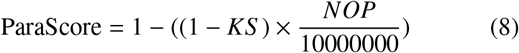

Table. 1 shows the ParaScore for each model along with the number of parameters and Kaggle Score. Kaggle Score which is 0.4×Dice coefficient + 0.6×Hausdorff distance is calculated by kaggle at the time of submission. The ParaScore of LWU-Net is the highest with 0.892 which suggests that the trade off between dice coefficient and the number of parameters is exceptional.

**Table 1:** Computational efficiency comparison LWU-Net and other traditional models. The table shows the, number of trainable parameters (NOP) for each model.

| Model | Number of parameters | Kaggle Score (KS) | ParaScore |
| --- | --- | --- | --- |
| U-Net | 31,043,651 | 0.82798 | 0.466 |
| U-Net + ResNet34 | 24,456,444 | 0.83434 | 0.595 |
| Dense U-Net | 17,004,899 | 0.65351 | 0.411 |
| U-Net + ResNet18 | 14,340,860 | 0.82892 | 0.755 |
| LWU-Net | 5,994,334 | 0.82037 | 0.892 |

**Figure 4:**
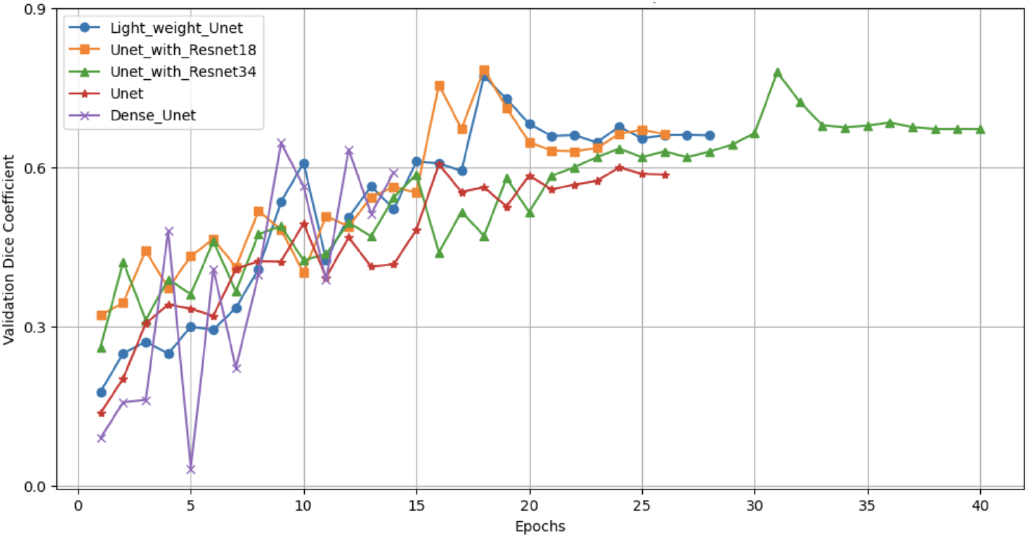
The plot of validation dice coefficients of different models versus the epoch number

Despite attempting various augmentation techniques such as ElasticTransform and ShiftScaleRotate, no noticeable improvement in the model’s performance was observed. Removing the rows with no segmentation from the dataset resulted in way too many false positives

The Adam optimizer was employed with a learning rate of 0.01. As shown in Fig. 6 the Light weight U-Net model was trained for 28 epochs, 18th epoch demonstrated the highest performance.

**Figure 5:**
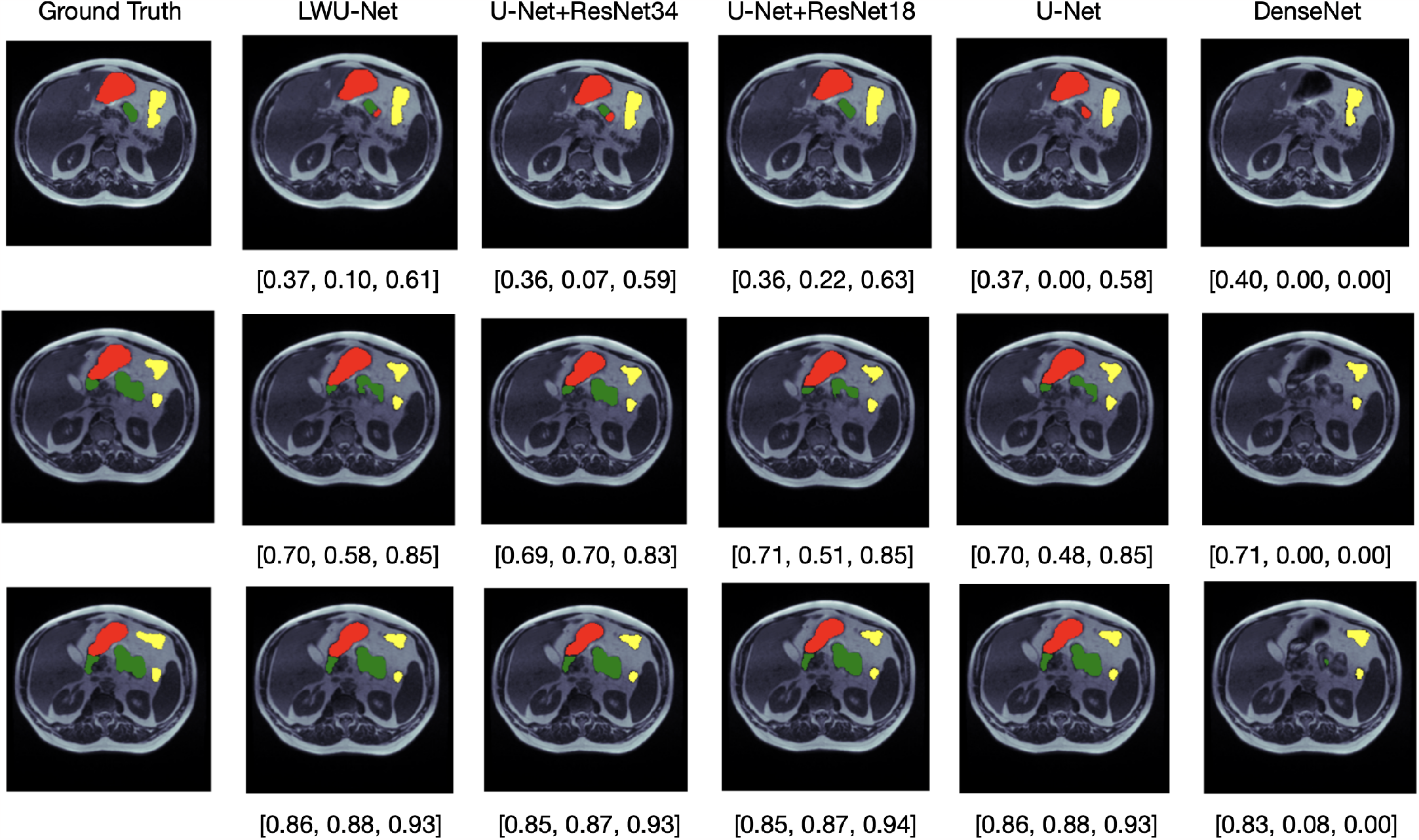
The visualization showcases the ground truth segmentation alongside its corresponding predicted segmentation mask predicted by different models along with the dice coefficients for large bowel, small bowel and stomach respectively, where red portion represents stomach, green represents small bowel and yellow represents large bowel

**Figure 6:**
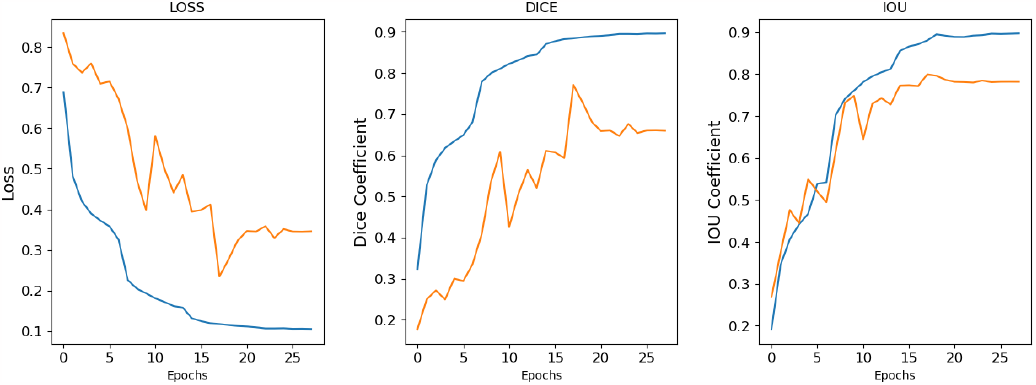
Plots illustrating the variation of the loss function, Dice coefficient, and Jaccard coefficient values with respect to the number of epochs during the training process of Light Weight U-Net.

As shown in the Table. 2 LWU-Net outperformed U-Net in both validation dice coefficient and validation IoU coefficient, the U-Nets with ResNet as baskbones have superior validation IoU coefficient but the validation dice coefficients are almost comparable with that of LWU-Net. Fig. 7 shows a plot comparing the kaggle score with respect to the number of parameters. Fig. 5 visualises the ground truth segmentation mask overlay and the predicted segmentation overlay generated by the mentioned models in Table. 2 from the validation set. From the comparison shown in Fig. 5 it is evident that LWU-Net model struggles in the segmentation of small bowel compared to the U-Net with ResNet34. The segmentation of large bowel is of the most accurate segmentation by all the models, later comes stomach with some minor differences but the the deciding segmentation is the small bowel.

**Table 2:** Performance comparison of LWU-Net with other traditional models. The table shows the, validation Dice coefficient, validation IoU coefficient for each model.

| Model | Validation Dice Coefficient | Validation IoU Coefficient |
| --- | --- | --- |
| U-Net | 0.6057 | 0.7755 |
| Dense U-Net | 0.6461 | 0.6234 |
| U-Net with ResNet18 | 0.7844 | 0.8218 |
| U-Net with ResNet34 | 0.7791 | 0.8269 |
| Light Weight U-Net | 0.7713 | 0.7993 |

**Figure 7:**
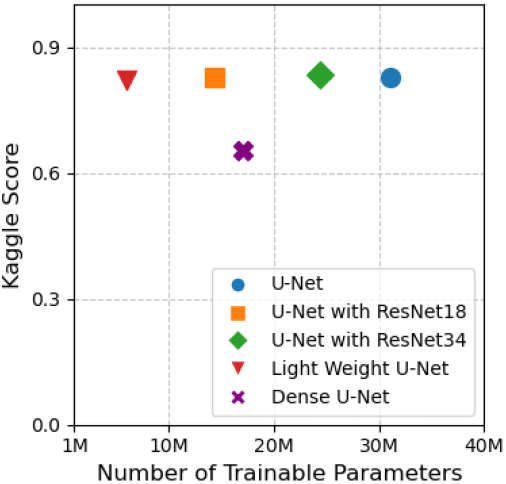
Plot illustrating the variation of kaggle score with respect to number of parameters of different models

## 5. Discussion

In Table 1, the LWU-net Model exhibits a significant reduction in complexity, having approximately 20% of the trainable parameters of the U-Net, about 25% of the U-Net equipped with a ResNet34 backbone, and nearly 40% of the U-Net with a ResNet18 backbone. Despite this substantial reduction in trainable parameters, the LWU-net model demonstrates only a marginal decrease in performance. This suggests that the LWU-net Model offers a commendable equilibrium between computational efficiency and segmentation accuracy.

## 6. Conclusion

The proposed LWU-Net architecture incorporating depthwise separable convolutions exhibits promising potential for achieving comparable segmentation performance with a substantial reduction in parameter count. This innovative approach proves to be valuable in image segmentation by striking a balance between model complexity and performance. It leads to efficient and resource friendly solutions. This reduction makes the architecture suitable for deployment with limited resources. The successful combination of design principles with the established U-Net framework opens up new possibilities for various applications, such as medical imaging and computer vision tasks. Therefore this advancement in network architecture represents a contribution to the on-going development of efficient and effective segmentation models, in the field of deep learning.

## Data Availability

Data is publicly accessible

https://kaggle.com/competitions/uw-madison-gi-tract-image-segmentation

